# In schizophrenia, the effects of the IL-6/IL-23/Th17 axis on health-related quality of life and disabilities are partly mediated by generalized cognitive decline and the symptomatome

**DOI:** 10.1101/2022.04.30.22274531

**Authors:** Ali Fattah Al-Musawi, Hussein Kadhem Al-Hakeim, Inas H. Al-Haboby, Abbas F. Almulla, Michael Maes

**Affiliations:** Department of Clinical Pharmacy and Laboratory Sciences, College of Pharmacy, University of Al-Kafeel, Iraq; Department of Chemistry, College of Science, University of Kufa, Iraq; Al-Diwaniya teaching hospital, Al-Diwaniya, Iraq; Medical Laboratory Technology Department, College of Medical Technology, The Islamic University, Najaf, Iraq; Department of Psychiatry, Faculty of Medicine, Chulalongkorn University, PathumWan, Bangkok, 10330, Thailand; Department of Psychiatry, Medical University of Plovdiv, Plovdiv, 4000, Bulgaria; Deakin University, IMPACT - the Institute for Mental and Physical Health and Clinical Translation, School of Medicine, Barwon Health, Geelong, Australia

**Keywords:** neuro-immune, inflammation, tryptophan catabolites, deficit schizophrenia, biomarkers, psychiatry

## Abstract

Schizophrenia patients show increased disabilities and lower quality of life (DisQoL). Nevertheless, there are no data whether, in schizophrenia, activation of the interleukin (IL)-6, IL-23, T helper (Th)-17 axis and lowered magnesium and calcium levels impact DisQoL scores. This study recruited 90 patients with schizophrenia (including 40 with deficit schizophrenia) and 40 healthy controls and assessed the World Health Association QoL instrument-Abbreviated version and Sheehan Disability scale, Brief Assessment of Cognition in Schizophrenia (BACS), IL-6, IL-23, IL-17, IL-21, IL-22, tumor necrosis factor (TNF)-α, magnesium and calcium. Regression analyses showed that a large part of the first factor extracted from the physical, psychological, social and environmental HR-QoL and interference with school/work, social life, and home responsibilities was predicted by a generalized cognitive deterioration (G-CoDe) index (a latent vector extracted from BACs scores), and the first vector extracted from various symptom domains (“symptomatome”), whereas the biomarkers had no effects. Partial Least Squares analysis showed that the IL6IL23Th17 axis and magnesium/calcium had highly significant total (indirect + direct) effects on HR-QoL/disabilities which were mediated by G-CoDe and the symptomatome (a first factor extracted from negative and positive symptoms). The IL6IL23Th17 axis explained 63.1% of the variance in a single latent trait extracted from G-CoDe, symptomatome, HR-QoL and disability data. The latter features are manifestations of a common core, namely the behavioral-cognitive-psycho-social worsening index, which is caused by the neuroimmunotoxic effects of the IL6IL23Th17 axis in subjects with lowered antioxidant defenses (magnesium and calcium) thereby producing damage to neuronal circuits underpinning deficit schizophrenia.

## Introduction

Schizophrenia is one of the most serious and debilitating mental illnesses, with an estimated worldwide prevalence of 0.32% (WHO, 2022). Personal, familial, social, educational, occupational, and other vital aspects of life are regularly affected by schizophrenia (WHO, 2022). Schizophrenia patients frequently report lowered health-related quality of life (HR-QoL) and increased functional limitations and disabilities in everyday living (Desalegn et al., 2020; Bobes et al., 2007; Kanchanatawan et al., 2019). There is now some agreement that assessing schizophrenia symptoms alone is inadequate for assessing schizophrenia’s intermediate and distal outcomes (Galuppi et al., 2010; Bobes et al., 2007). HR-QoL and social functioning are now well-established distal outcome measures of schizophrenia that are highly valued by patients and their clinicians (Galuppi et al., 2010; Bobes et al., 2007).

Nevertheless, in schizophrenia, HR-QoL may be impacted by psychiatric symptoms such as psychosis, hostility, excitation, mannerism (PHEM) and negative (blunting of affect, alogia, avolition, anhedonia) symptoms (Kanchanatawan et al., 2019; Galuppi et al., 2010; Sim et al., 2004; Ritsner et al., 2005; Becker et al., 2005; Fitzgerald et al., 2001; Gorna et al., 2014; Savill et al., 2016). There is some debate as to whether negative, positive, or affective symptoms predict lowered HR-QoL in schizophrenia (Kanchanatawan et al., 2019). Nevertheless, recently we have shown that one latent construct could be extracted from PHEM) and negative (PHEMN) symptoms and that this common core predicts HR-QoL (Kanchanatawan et al., 2019; Maes and Kanchanatawan, 2021). These results indicate that a generalized index of symptom severity (labeled as the “symptomatome”), rather than a specific symptom domain, largely determines lowered HR-QoL in schizophrenia (Maes and Kanchanatawan et al., 2021). Moreover, HR-QoL is significantly lower in deficit as compared with nondeficit schizophrenia (Sum et al., 2018; Kanchanatawan et al., 2019), which may be explained by findings that the former subtype is characterized by increased severity of PHEMN symptoms (Kanchanatawan et al., 2019).

Not only the symptomatome of schizophrenia may have a negative effect on HR-QoL, but also the cognitive deficits associated with schizophrenia might have a negative effect in such individuals (Ueoka et al., 2011; Alptekin et al., 2005; Tolman and Kurtz, 2012; Mohamed et al., 2008; Kanchanatawan et al., 2019). A meta-analysis revealed significant relationships between objective HR-QoL and cognitive deficits such as working memory, executive functioning, fluency, list learning, and processing speed, but with a small impact size (Tolman and Kurtz, 2012). Ueoka et al. (2011) found relationships with attention, verbal memory, and information processing speed, while Alptekin et al. (2005) found links with executive dysfunction and working memory deficiencies. Additionally, Keefe and Harvey (2012) observed that schizophrenia’s cognitive deficiencies impair social functioning, including the ability to work, get employment, live independently, and function properly, and Mohamed et al. (2008) reported that negative and positive symptoms contribute more to decreased HR-QoL than cognitive deficiencies. Nonetheless, we recently discovered that a common core of cognitive deficits across multiple domains, including episodic, semantic, and working memory, executive functions, and attention, was strongly associated with HR-QoL in schizophrenia, and that these effects were partially mediated by the symptomatome (Maes and Kanchanatawan, 2021). As such, this generalized cognitive decline (G-CoDe) is a critical element in explaining decreased HR-QoL in schizophrenia patients.

A recent study established that a one latent construct underpins the G-Code’s cognitive domains, PHEMN symptoms as well as the physical, psychological, social, and environmental subdomain scores of the HR-QoL (Maes and Kanchanatawan, 2021). As a result, this common core integrates behavioral, cognitive, physical, and psychosocial (BCPS) components into a single validated underlying factor that represents BCPS problems and so serves as an overall worsening measure of schizophrenia (Maes and Anderson, 2021; Maes et al., 2021b).

Additionally, there is now evidence that first episode psychosis and schizophrenia (FEP/FES) as well as deficit schizophrenia are systemic neuroimmunotoxic illnesses that may develop in response to immunological stimuli in patients who have impaired immune regulation and neuroprotection (Smith and Maes, 1995; Roomruangwong et al., 2020; Maes et al., 2021a; 2021b). These schizophrenia phenotypes are characterized by increased neuroimmunotoxic biomarkers, including interleukin (IL)-1, IL-6, IL-17, tumor necrosis factor (TNF)-α, and eotaxin (CCL-11) (Roomruangwong et al., 2020; Maes et al., 2021b), and activation of the tryptophan catabolite (TRYCAT) pathway with increased levels of IgA directed to neurotoxic TRYCATs such as picolinic (PA), xanthurenic (XA) and quinolinic acid (QA) and 3-OH-kynurenine (3OHK) (Roomruangwong et al., 2020; Maes et al., 2021b). According to the neuroimmunotoxic theory of schizophrenia, these (and other) neuroimmunotoxic pathways and underlying intracellular networks may affect functional connectome circuits in specific brain areas, resulting in BCPS worsening as measured by the latent vector score underlying the G-CoDe, symptomatome, and HR-QoL values (Maes et al., 2021b; Maes and Anderson, 2021). In this regard, we established that a) the effects of immune activation on the symptomatome were mediated via the path from elevated levels of neuroimmunotoxic products (including TRYCATs) to the G-CoDe (Sirivichayakul et al., 2019); b) the effects of neuroimmunotoxic products on HR-Qol are mediated via the G-CoDe and its effects on the symptomatome (Maes and Kanchanatawan, 2021); and c) a larger part of the variance in the BCPS-worsening index is explained by effects of diverse neuroimmunotoxic compounds (Maes et al., 2021b).

We recently created a novel neuroimmunotoxic pathway index in schizophrenia, particularly deficit schizophrenia, by connecting IL-6, IL-23, and IL-17 to critical actors such as IL-21, IL-22, and TNF-α (Al-Hakeim et al., 2022). As previously stated, these cytokines all show neuroimmunotoxic properties and, more crucially, are associated with the cognitive deficits and symptomatome of schizophrenia (Al-Hakeim et al., 2022). In addition, one validated latent vector could be extracted from these cytokines, validating the novel concept of the “neuroimmunotoxic IL6IL23Th17 axis” (Al-Hakeim et al., 2022).

Nevertheless, there are no data whether this axis affects HR-QoL and disabilities in schizophrenia and whether these effects are mediated via the G-CoDe or symptomatome. Hence, the current study was performed to examine whether the effects of the IL6IL23Th17 axis on HR-Qol and dysfunctions in community living are mediated by the G-CoDe and or the symptomatome.

## Subjects and Methods

### Participants

This research included 90 individuals with schizophrenia and 40 healthy controls. Between February and June 2021, patients were recruited at the psychiatry unit of Al-Hakeem General Hospital in Najaf Governorate, Iraq. Controls were family and friends of staff and friends of patients and they were recruited from the same catchment area as the patients (Najaf Governorate). All schizophrenia patients were diagnosed using DSM-IV-TR criteria using the Mini*-*International Neuropsychiatric Interview (M.I.N.I.). All patients had been stable for at least 12 weeks and they were divided into these with and without deficit schizophrenia as diagnosed using the Schedule for the Deficit Syndrome (SDS) (Kirkpatrick et al., 1989). Deficit schizophrenia was defined as the presence of at least two of the following six symptoms with clinically significant severity during the preceding 12 months: restricted affect, diminished emotional range, poverty of speech, curbing of interest, diminished sense of purpose, and diminished social drive. Additionally, to be classified as primary deficit schizophrenia, the symptoms must not be related to antipsychotic medication-induced extrapyramidal side effects.

Patients with axis-1 DSM-IV-TR disorders were excluded, including schizoaffective disorder, major depression, bipolar disorder, obsessive-compulsive disorder, substance use disorders, and psycho-organic diseases. None of the controls had ever been diagnosed with DSM-IV-TR axis 1 disorders. Patients and controls were excluded if they had been diagnosed with a neurodegenerative or neuroinflammatory condition such as Alzheimer’s disease, Parkinson’s disease, multiple sclerosis, or stroke; if they had ever had immunosuppressive or glucocorticoid therapy, if they suffered from (auto)immune disorders such as chronic obstructive pulmonary disease, inflammatory bowel disease, psoriasis, rheumatoid arthritis, or diabetes mellitus type 1, or if they had taken antioxidant supplements (n3 poly-unsaturated fatty acids, N-acetyl cysteine) in therapeutic dosages. We also excluded pregnant or lactating women.

The research was conducted in accordance with the Iraqi and international privacy and ethical legislation. All participants and first-degree relatives of patients with schizophrenia provided written informed permission before participation in this research (legal representatives are mother, father, brother, spouse, or son). The ethics commission (IRB) of the College of Science, University of Kufa, Iraq (82/2020) authorized the research, which adheres to the Declaration of Helsinki’s International Guideline for the Protection of Human Subjects.

### Clinical assessments

Semi-structured interviews with patients and controls were done by a senior psychiatrist with experience in schizophrenia. The same psychiatrist evaluated the M.I.N.I., to make the diagnosis of schizophrenia, and the SDS to make the diagnosis of primary deficit schizophrenia. On the same day as the semistructured interview, we assessed HR-QoL using the WHO-QoL-BREF (World Health Organization Quality of Life Instrument-Abridged Version) (WHO, 1993). This scale rates 26 items across four categories of HR-QoL: 1) Domain 1 or physical health: energy, sleep, fatigue, pain, discomfort, work capacity, activities of daily living, medication dependence, and mobility; 2) Domain 2 or psychological health: self-esteem, body image, learning, thinking, concentration, memory, negative and positive feelings, and beliefs (spirituality-religion-personal); 3) Domain 3 or social relationships: social support, sexual activity, and personal relationships; and 4) Domain 4 or environment: physical safety and security. We calculated the raw scores for the four domains using the WHO-QoL-BREF criteria (WHO, 1993). The Sheehan scale (Sheehan and Sheehan, 2008) was used to assess disabilities, which is a self-administered rating scale with scores ranging from 0 to 10 for domains such as family, occupational, and social, activities, with a score of 0-3 indicating mild disabilities, 4-6 indicating moderate disabilities, and 7-10 indicating severe disabilities. We next derived the first factor from the raw HR-QoL and Sheehan domain scores, named the “DisQoL phenomenome”.

The same day, the senior psychiatrist also measured the Scale for the Assessments of Negative Symptoms (SANS) (Andreasen, 1989), the Brief Psychiatric Rating Scale (BPRS) (Overall and Gorham, 1962), the Positive and Negative Syndrome Scale (PANSS) (Kay et al., 1987), and the Hamilton Depression (HAM-D) and Anxiety (HAM-A) Rating Scales (Hamilton, 1959; 1960). We constructed z-unit weighted composite scores indicating psychosis, hostility, excitement, mannerism, psychomotor retardation (PMR), and formal thought disorders (FTD) using BPRS, HAM-D, HAM-A, and PANSS items, as previously described, and the total SANS, negative PANSS symptom sore and the total SDS score were used to quantify negative symptoms (Maes et al., 2021b). We next derived the first factor extracted from the PHEMN, PMR and FTD domains, dubbed the “symptomatome”.

On the same day, a research psychologist conducted neuropsychological tests using the Brief Assessment of Cognition in Schizophrenia (BACS) (Keefe et al., 2004) while remaining blind to the clinical diagnosis. The latter battery consists of the List Learning Test (which measures verbal episodic memory), Digit Sequencing Task (which measures working memory), Category Instances (which measures semantic fluency), the Controlled Word Association (which measures letter fluency), Symbol Coding (which measures attention), the Tower of London (which measures executive functions), and the token motor task. Consequently, we extracted the first factor from the six BACS test results (not the token motor task) and refer to it as the “G-CoDe” (generalized cognitive decline) (Maes and Kanchanatawan, 2021). Tobacco use disorder (TUD) was diagnosed in accordance with DSM-IV-TR criteria. Body mass index (BMI) was computed using body weight in kilograms (kg) / length in meters (m)^2^.

### Assays

Fasting venous blood samples were taken from each patient in the early morning hours. Following 15 minutes at room temperature, blood was allowed to coagulate for 10 minutes before being centrifuged for 10 minutes at 3000 rpm. After serum separation, it was transferred to Eppendorf tubes and kept at -80 °C until the assays were caried out. Serum IL-6, IL-10 and IL-1β levels were determined using commercial ELISA sandwich kits purchased from Sunlong Biotech Co., Ltd (Zhejiang, China), and Melsin Medical Co. (Jilin, China) provided the remaining ELISA kits (IL-17, IL-21, IL-22, IL-23, and TNF-α). The kits detected IL-10, IL-17, IL-21, IL-22, IL-23, and TNF-α at 1.0 pg/ml and IL-1β and IL-6 at 0.1 pg/ml. Calcium and magnesium were measured using spectrophotometric kits supplied by Biolabo® (Maizy, France). The coefficient of variance within tests was <10.0% for all assays (precision within-assay).

### Statistical analysis

To compare scale variables across groups, one-way analysis of variance (ANOVA) or the Kruskal-Wallis test were performed, while category variables were analyzed using analysis of contingency tables (χ2-tests). Multiple regression analysis (automatic technique with a p-to-entry of 0.05 and a p-to-remove of 0.06 while evaluating the change in R2) was used to identify the critical explanatory variables (e.g. biomarkers, G-CoDe or phenome data) that predict dependent variables (e.g. the worsening index) in all individuals combined or in a restricted study sample comprising schizophrenia patients only. Homoscedasticity was checked using the White and modified Breusch-Pagan tests for homoscedasticity; multicollinearity was determined using tolerance and VIF, and multivariate normality was determined using Cook’s distance. The results of regression analyses were always bootstrapped using 5.000 bootstrap samples, and the latter are presented if the findings were not concordant. The associations among study groups based on HR-QoL and disabilities and biomarkers, G-Code or phenome data were examined using multivariate GLM analysis with age, gender, education, BMI, TUD, and/or the drug state as covariates. Consequently, we determined the model’s estimated marginal mean values (SE), which are the marginal means adjusted for the confounders. Multiple pair-wise differences were assessed using protected Least Significant Difference (LSD) testing. Multiple associations were submitted to p-correction for false discovery rate (FDR) (Benjamini and Hochberg, 1995). Factor analysis unweighted least squares extraction was performed to delineate factors reflecting latent constructs. The Kaiser-Meyer-Olkin measure of sampling adequacy was used to check factoriability. Factors were considered appropriate when all loadings were >0.6, the explained variance > 50.0% and when Cronbach alpha was > 0.7. All statistical analyses were conducted using IBM SPSS Windows version 28.

Partial Least Squares (PLS)-SEM pathway analysis is a statistical technique for predicting complex cause-effect relationships by combining single indicators and latent variables (factors based on a set of strongly related indicators) (Ringle et al., 2014). PLS enables estimation of multi-step mediation models with multiple latent components, single indicator variables, and structural pathways without imposing distributional assumptions on the data. By merging the several components of an illness into a causal-effect, mediation model, this method has recently been applied to build innovative nomothetic models of affective disorders and schizophrenia (Sirivichayakul et al., 2019). A causal framework is constructed using the causome, protectome, adverse outcome pathways, and phenome indicators (single or latent vectors), which is then evaluated and cross-validated using PLS pathway analysis on bootstrapped data. Complete pathway analysis is undertaken using 5000 bootstrap samples only when the outer and inner models met the following quality criteria: a) the SRMR index is less than 0.08; b) the latent vectors have adequate construct validity as judged by average variance extracted > 0.5, rho A > 0.7, composite reliability > 0.8, and Cronbach’s alpha > 0.7; and c) the outer model loadings on the latent vectors are more than 0.6 at p <0.001. The model’s prediction performance was assessed using PLSpredict and a 10-fold cross-validation procedure. We explored compositional invariance utilizing Predicted-Oriented Segmentation analysis, Multi-Group Analysis, and Measurement Invariance Assessment. Subsequently, pathway coefficients (with precise p values), specific indirect (mediated) effects, and total (overall) effects are assessed to ascertain the direct and mediated pathways’ impact. According to G Power analysis, the anticipated sample size for a multiple regression analysis (which is relevant to PLS) should be at least 70 to achieve 0.8 power, 0.2 effect size, and 0.05 alpha with five predefined variables.

## Results

### Socio-demographic data

In order to display the demographic, clinical, and biomarker data is relation to the HR-QoL/disability data we extracted the first factor (unweighted least squares extraction) from the 3 Sheehan domains and 4 HR-QoL domains (labeled “DisQoL” factor; KMO=0.913, the first factor explained 73.5% of the variance and all 7 loadings were > 0.714) and we subjected this DisQoL phenomenome factor score to a visual binning method which allowed to divide the sample into three groups using cut-off values of -0.6 and +0.4, namely those with normal DisQol scores (all controls and 2 schizophrenia patients) and patients with moderate and severe DisQoL values.

**Table 1** shows the demographic features of these 3 subgroups. There were no significant differences in age, BMI, sex and urban/rural distribution, marital status, TUD, and employment status between the three subgroups. Education was somewhat lower in the moderate and severe DisQoL groups than in the normal DisQoL group. The duration of illness was significantly greater and age of onset lower in those with severe DisQoL scores as compared with those with less severe DisQoL scores. Table 1 shows that the 4 HR-QoL and 3 disability subdomain scores were significantly different between the three groups. There were no significant differences in the treatments between the severe and moderate groups except for olanzapine which was marginally higher in the moderate DisQoL group (without FDR p correction).

**Table 1.** Sociodemographic and clinical variables in healthy controls and schizophrenia patients divided into subjects with severe disabilities and low quality of life (severe DisQoL), moderate changes in disabilities and DisQoL (moderate DisQoL) and those with normal DisQoL values (normal DisQoL)

| Parameter | Normal DisQoL<sup>A</sup><br>n=42 | Moderate DISQoL<sup>B</sup><br>n=52 | Severe DISQoL<sup>C</sup><br>n=36 | F/$\chi^2$ | df | p |
| --- | --- | --- | --- | --- | --- | --- |
| Age (years) | 36.1 ± 12.0 | 38.3 ± 12.6 | 36.9 ± 14.5 | 0.35 | 2/127 | 0.706 |
| Female /Male ratio | 21/21 | 21/31 | 15/21 | 0.97 | 2 | 0.628 |
| BMI (kg/m<sup>2</sup>) | 26.82 ± 3.49 | 27.07 ± 3.77 | 27.00 ± 4.09 | 0.05 | 2/127 | 0.950 |
| Education Year | 12.1 ± 3.2<sup>B,C</sup> | 9.1 ± 3.7<sup>A,C</sup> | 9.5 ± 3.3<sup>A,B</sup> | 9.99 | 2/127 | <0.001 |
| Married/Single | 19/23 | 21/31 | 15/21 | 0.23 | 2 | 0.901 |
| Rural/Urban | 17/ 25 | 19/33 | 14/22 | 0.16 | 2 | 0.973 |
| TUD (No /Yes) | 26/16 | 27/ 25 | 24/12 | 2.10 | 2 | 0.375 |
| Employment (No/Yes) | 15/ 27 | 18/ 34 | 19/17 | 7.16 | 4 | 0.109 |
| Family history No/Yes | 41/1 | 34/18 | 17/19 | 25.02 | 2 | <0.001 |
| HC / NonDef SCZ/ Def SCZ | 40 / 2 / 0 | 0/ 42/ 10 | 0 / 1 / 35 | 196.023 | 4 | <0.001 |
| Duration of Disease (years) | - | 8.4 ± 7.6<sup>C</sup> | 13.0 ± 8.6<sup>B</sup> | 7.01 | 1/86 | 0.010 |
| Age of onset (years) | - | 29.9 ± 9.9<sup>C</sup> | 23.6 ± 11.9<sup>B</sup> | 7.32 | 1/86 | 0.008 |
| Sheehan, Work-School | 1.26 ± 0.94<sup>B,C</sup> | 5.42 ± 1.46<sup>A,C</sup> | 7.00 ± 1.69<sup>A,B</sup> | 184.07 | 2/127 | <0.001 |
| Sheehan, Social life | 1.00 ± 0.77<sup>B,C</sup> | 4.54 ± 1.41<sup>A,C</sup> | 7.92 ± 1.27<sup>A,B</sup> | 325.45 | 2/127 | <0.001 |
| Sheehan, Family life | 1.24 ± 0.69<sup>B,C</sup> | 4.35 ± 1.40<sup>A,C</sup> | 7.39 ± 1.15<sup>A,B</sup> | 281.49 | 2/127 | <0.001 |
| WHO-QoL, physical | 29.1 ± 3.3<sup>B,C</sup> | 20.6 ± 3.3<sup>A,C</sup> | 14.8 ± 3.4<sup>A,B</sup> | 186.68 | 2/127 | <0.001 |
| WHO-QoL, psychological | 24.2 ± 3.0<sup>B,C</sup> | 17.3 ± 2.2<sup>A,C</sup> | 13.2 ± 3.3<sup>A,B</sup> | 159.24 | 2/127 | <0.001 |
| WHO-QoL, social | 10.9 ± 1.8<sup>B,C</sup> | 8.1 ± 1.9<sup>A,C</sup> | 6.9 ± 2.0<sup>A,B</sup> | 45.36 | 2/127 | <0.001 |
| WHO-QoL, environment | 30.5 ± 3.3<sup>B,C</sup> | 24.9 ± 3.2<sup>A,C</sup> | 22.9 ± 3.9<sup>A,B</sup> | 52.60 | 2/127 | <0.001 |
| Benzodiazepines No/Yes | 42 / 0 | 43 / 9 | 26 / 10 | 1.37* | 1 | 0.296 |
| Mood stabilizers No/Yes | 42 / 0 | 32 / 20 | 17 / 19 | 1.77* | 1 | 0.199 |
| Olanzapine No/Yes | 41 / 1 | 11 / 41 | 16 / 20 | 5.43* | 1 | 0.020 |
| Clozapine No/Yes | 42 / 0 | 48 / 4 | 28 / 8 | 3.81* | 1 | 0.051 |
| Fluphenazine No/Yes | 42 / 0 | 37 / 15 | 24 / 12 | 0.20* | 1 | 0.814 |
| Haloperidol No/Yes | 42 / 0 | 50 / 2 | 31 / 5 | FEPT* | - | 0.117 |
| Quetiapine No/Yes | 42 / 0 | 49 / 3 | 34 / 2 | FEPT* | - | 1.0 |
| Risperidone No/Yes | 42 / 0 | 45 / 7 | 27 / 9 | 1.90* | 1 | 0.260 |
| Fluoxetine No/Yes | 42 / 0 | 50 / 2 | 35 / 1 | FEPT* | - | 1.0 |

Data are shown as mean (SD) or as ratios. F: results of analysis of variance; $\chi^2$ : results of analysis of contingency tables; FEPT: Fisher's exact probability test. <sup>A, B, C</sup>: Pairwise comparison among group means.\*: these analyses compare the moderate and severe DisQoL groups.
TUD: tobacco use disorder; NonDef SCZ/ Def SCZ: non-deficit and deficit schizophrenia.

The table also shows that there was a significant association between the DisQol groups and the classification into controls, deficit and non-deficit schizophrenia, with most deficit patients (35/45) being allocated to the severe DisQol group while only 1 out of 43 non-deficit schizophrenia patients was allocated to the severe DisQol group.

### Differences in clinical ratings between the three DisQoL groups

**Table 2** shows the results of ANOVAs which examine the associations between the DisQoL groups and clinical data including the symptomatome and cognitome data. We used the Kruskal-Wallis test because there was heterogeneity of variance for some variables between the groups. After FDR p correction we found that all comparisons were significant using an FDR of p=0.05. All PHEMN symptom domains increased from the normal → moderate → severe DisQoL group, while the cognitive test results decreased from the normal → moderate → severe DisQoL group. We also used univariate GLM analyses (on the Log transformations) with sex, age and education as covariates and found that the latter did not have significant effects.

**Table 2.**
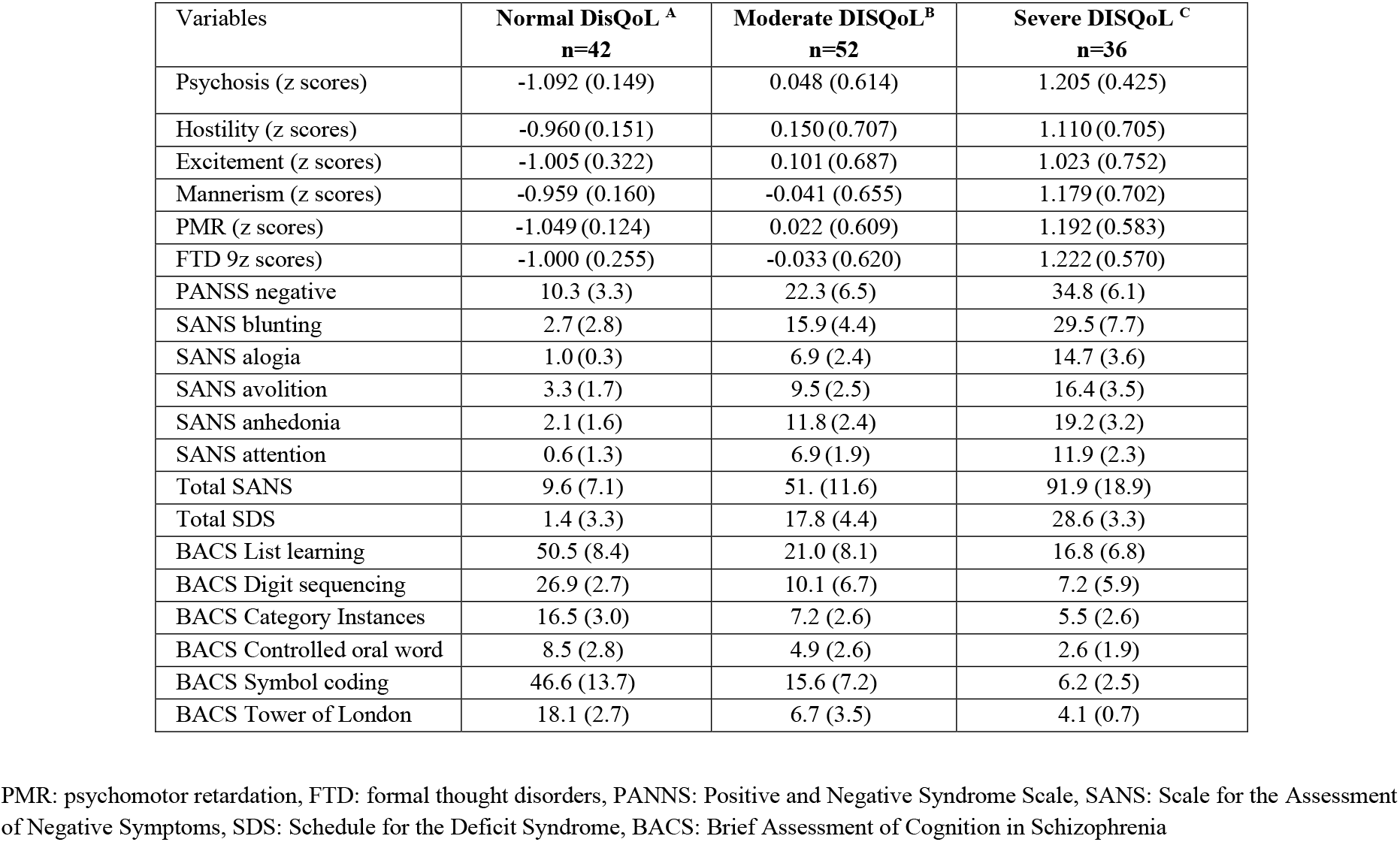
Clinical data in healthy controls and schizophrenia patients divided into subjects with severe disabilities and low quality of life (severe DisQoL), moderate changes in disabilities and DisQoL (moderate DisQoL) and those with normal DisQoL values (normal DisQoL)

### Differences in biomarkers between the three DisQoL groups

**Table 3** examines the associations between the DisQol groups and all 10 biomarkers, namely magnesium, calcium, the 8 cytokines and two cytokine profiles, namely a) a composite score computed as z score IL-6 (z IL-6) + z IL-17 + z IL-23 indicating pathogenic Th17 signaling, and b) a latent vector (LV) extracted from IL-17, IL-21, IL-22, and TNF-α (AVE=58.0%, composite reliability=0.845, all loadings > 0.620) and IL-6 and IL-23 (AVE=58.7%, composite reliability=0.738, loadings: 0.823 and 0.705, respectively). This construct was dubbed the IL6IL23Th17 axis (Hussein et al., 2022) and showed adequate quality data, namely AVE=70.8%, loadings: > 0.841 and composite reliability=0.844). The results of univariate GLM analyses with age, sex, smoking and BMI as covariates, showed that the biomarkers were significantly different between the DisQoL groups with lowered magnesium and calcium and increased IL-6, IL-21, IL-17, and pathogenic Th17 and IL6IL23Th17 axis indices in the severe DisQoL group as compared with the moderate and normal DisQoL groups. IL-1β, IL-23 IL-22, TNF-α and IL-10 were significantly higher in the severe/moderate than in the normal DisQoL groups.

**Table 3.** Biomarker data in healthy controls and schizophrenia patients divided into subjects with severe disabilities and low quality of life (severe DisQoL), moderate changes in disabilities and DisQoL (moderate DisQoL) and those with normal DisQoL values (normal DisQoL)

| **Biomarkers** | **Normal DisQoL<sup>A</sup>** | **Moderate DisQoL<sup>B</sup>** | **Severe DisQoL<sup>C</sup>** | **F (df=2/122)** | **p** |
| --- | --- | --- | --- | --- | --- |
| Magnesium (mM) | 1.008 (0.021)<sup>C</sup> | 0.989 (0.019)<sup>C</sup> | 0.899 (0.022)<sup>A, B</sup> | 6.97 | 0.001 |
| Calcium (mM) | 2.383 (0.042)<sup>C</sup> | 2.348 (0.037)<sup>C</sup> | 2.216 (0.045)<sup>A, B</sup> | 4.27 | 0.016 |
| IL-1β (pg/mL) | 2.46 (0.149)<sup>B, C</sup> | 3.50 (0.131)<sup>A</sup> | 3.80 (0.158)<sup>A</sup> | 21.27 | <0.001 |
| IL-6 (pg/mL) | 5.92 (0.57)<sup>C</sup> | 6.26 (0.50)<sup>C</sup> | 8.35 (0.60)<sup>A, B</sup> | 5.33 | 0.006 |
| IL-23 (pg/mL) | 14.73 (2.48)<sup>B, C</sup> | 24.63 (2.17)<sup>A</sup> | 27.02 (2.62)<sup>A</sup> | 6.61 | 0.002 |
| IL-22 (pg/mL) | 17.94 (1.19)<sup>B, C</sup> | 28.06 (1.04)<sup>A</sup> | 29.31 (1.26)<sup>A</sup> | 26.62 | <0.001 |
| IL-21 (pg/mL) | 159.88 (14.66)<sup>B, C</sup> | 232.93 (12.85)<sup>A, C</sup> | 279.40 (15.51)<sup>A, B</sup> | 15.90 | <0.001 |
| TNF-α (pg/mL) | 19.99 (2.14)<sup>B, C</sup> | 43.75 (1.88)<sup>A</sup> | 45.84 (2.26)<sup>A</sup> | 43.61 | <0.001 |
| IL-17 (pg/mL) | 17.56 (1.71)<sup>B, C</sup> | 43.28 (1.50)<sup>A, C</sup> | 47.87 (1.81)<sup>A, B</sup> | 87.67 | <0.001 |
| IL-10 (pg/mL) | 7.43 (0.63)<sup>B, C</sup> | 12.22 (0.55)<sup>A</sup> | 11.47 (0.66)<sup>A</sup> | 16.87 | <0.001 |
| Th17 pathogenic (z scores) | -0.860 (0.127)<sup>B, C</sup> | 0.215 (0.111)<sup>A, C</sup> | 0.692 (0.132)<sup>A, B</sup> | 36.47 | <0.001 |
| IL23IL16Th17 axis (z scores) | -0.973 (0.116)<sup>B, C</sup> | 0.271 (0.102)<sup>A, C</sup> | 0.744 (0.120)<sup>A, B</sup> | 54.37 | <0.001 |

IL: interleukin, TNF: tumor necrosis factor,
Results are shown as model (univariate GLM analysis)-generated marginal means (SE) after covarying for age, sex, tobacco use disorder and body mass index.

### Results of PLS-SEM path modeling

Figure 1. shows a first PLS model which examined the associations between the DisQoL phenomenome (a LV extracted from the 4 HR-QoL and 3 Sheehan domains) as final outcome variable, and input variables including education, magnesium, calcium, IL-6 and IL-23 (entered as single indicators), the Th17-TNF axis (a LV extracted from IL-17, IL-1β, IL-21, IL-22 and TNF-α), the G-CoDe (a LV extracted from the BACs test results), and the symptomatome (a LV extracted from the negative PANSS score, the total SDS score, the SANS items and total score, PHEM symptoms, PMR and FTD). The model quality was adequate with SRMR=0.034, and we detected adequate values for construct reliability validity of the DisQoL phenomenome (AVE=0.769; composite reliability=0.959, Cronbach alpha=0.949, all loadings > 0.762); symptomatome (AVE=0.897; composite reliability=0.992, Cronbach alpha=0.991, all loadings > 0.897); G-CoDe (AVE=0.806; composite reliability=0.961, Cronbach alpha=0.951, all loadings > 0.773); and Th17-TNF axis (AVE=0.546; composite reliability=0.856, Cronbach alpha=0.789, all loadings > 0.604). Blindfolding showed that the DisQoL phenomenome LV (0.708), symptomatome LV (0.673), G-CoDe LV (0.586), and Th17-TNF LV (0.090) showed adequate construct cross-validated redundancies. Figure 1 shows the results of complete PLS analysis (bias corrected and accelerated with 5000 bootstraps,): a large part of the variance in the DisQoL phenomenome LV was explained by the regression on the symptomatome and G-CoDe LV and the Th17-TNF axis, although the effects of the latter were minimal; 75.9% of the variance in the symptomatome LV was explained by the G-CoDe LV and calcium (both inversely), and the Th17-TNF LV and IL-6 (both positively); 73.6% of the variance in the G-CoDe LV was explained by education, magnesium and the Th17-TNF LV; and 17.6% of the variance in the latter LV was explained by the regression on IL-6 and IL-23. There were significant total effects of IL-6 (t=2.45, p=0.007), IL-23 (t=4.34, p<0.001) and the Th17-TNF LV (t=23.49, p<0.001) on the DisQoL phenomenome LV. The effects of IL-6 on the phenomenome were mediated (specific indirect effects) by the paths from Th17-TNF to the symptomatome (t=1.83, p=0.034), IL-23 to Th17-TNF to G-CoDe (t=1.91, p=0.028), Th17-TNF to G-CoDe (t=1.82, p=0.034), IL-23 to Th17-TNF to G-CoDe to symptomatome (t=1.99, p=0.034), IL-23 to Th17-TNF to symptomatome (t=1.91, p=0.028), and Th17-TNF to G-CoDe to symptomatome (t=1.95, p=0.026). The effects of IL-23 on the DisQoL phenomenome were mediated by Th17-TNF (t=1.76, p=0.040), and the paths from Th17-TNF to symptomatome LV (t=3.01, p=0.001), Th17-TNF to G-CoDe (t=3.50, p<0.001) and Th17-TNF to G-CoDe to symptomatome (t=3.81, p<0.001). The effects of the Th17-TNF LV on the DisQoL phenomenome were partially mediated by G-CoDe (t=5.77, p<0.001), symptomatome LV (t=4.21, p<0.001) and the path from G-CoDe to symptomatome (t=6.90, p<0.001). Part of the effects of the G-CoDe on the DisQoL phenomenome was mediated by symptomatome LV (t=6.99, p<0.001). **Figure 2 and 3** show the partial regressions of the DisQol phenomenome on the symptomatome and G-CoDe, respectively (after adjusting for the effects of age, sex, BMI, TUD, education, calcium, magnesium and the IL6IL23Th17 axis, which was not significant in this regression). These associations between the DisQoL score and the symptomatome (partial r=0.661, p<0.001, n=89) and G-CoDe (r=-0.224, p=0.043) were also significant in the restricted sample of schizophrenia patients.

**Figure 1.**
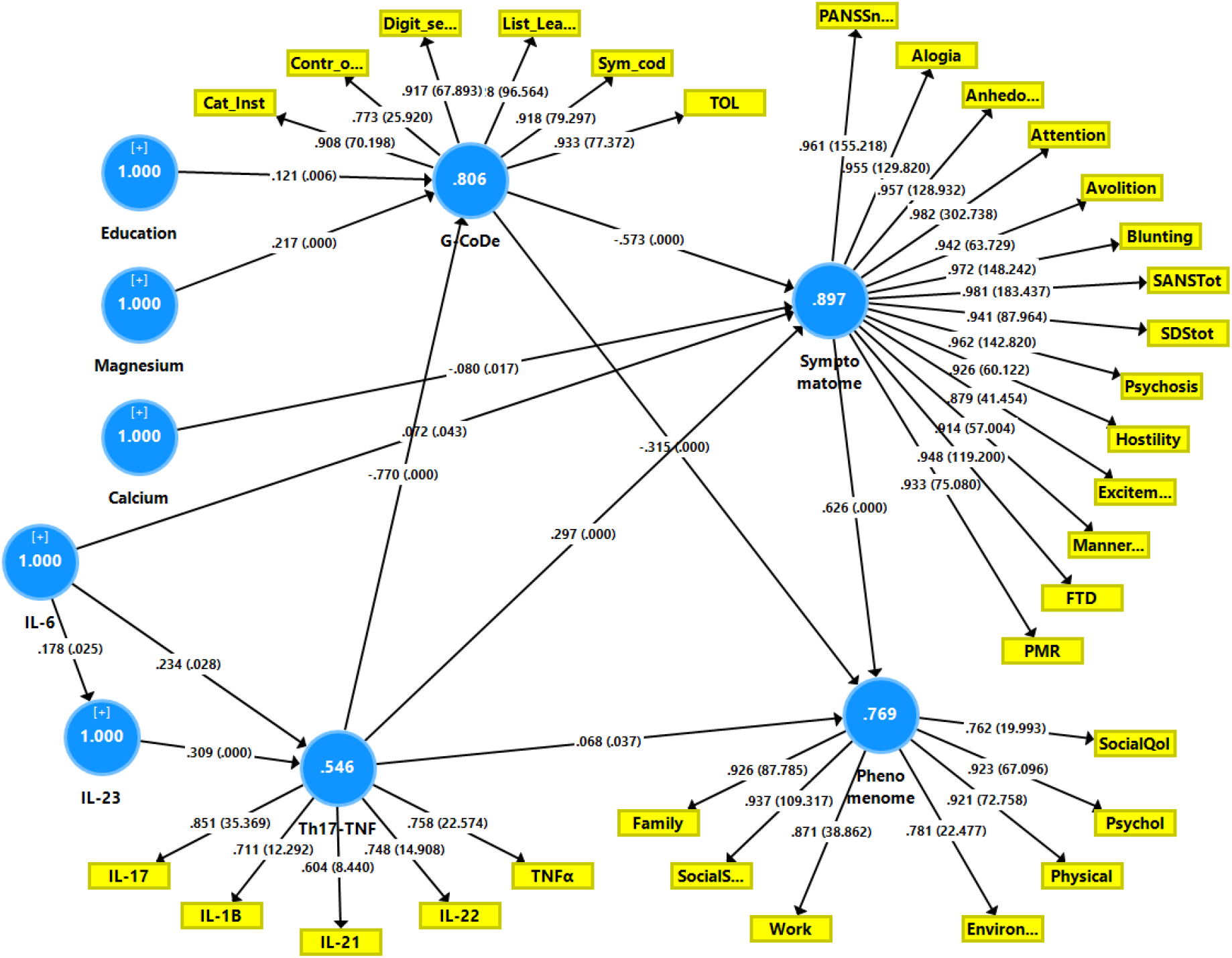
Results of Partial Least Squares (PLS) path analysis. This PLS analysis examines the associations between the DisQoL phenomenome, a latent vector (LV) extracted from 3 Sheehan disability (Dis) domains and 4 health related quality of Life (QoL) data as final outcome variable. Input variables include education, magnesium, calcium, interleukin (IL)-6 and IL-23 (entered as single indicators), the T helper (Th)17-TNF axis, a LV extracted from IL-17, IL-1β, IL-21, IL-22 and tumor necrosis factor (TNF)-α, the G-CoDe (a LV extracted from the cognitive test results) and the symptomatome (a LV extracted from the negative PANSS score, the total SDS score, the SANS items and total score and other symptoms, including psychomotor retardation (PMR) and formal thought disorders (FTD). PANNS: Positive and Negative Syndrome Scale, SANS: Scale for the Assessment of Negative Symptoms, SDS: Schedule for the Deficit Syndrome, BACS: Brief Assessment of Cognition in Schizophrenia Shown are path coefficients with p value, the factor loadings with p value and average variances explained (white figures in the blue circles).

**Figure 2.**
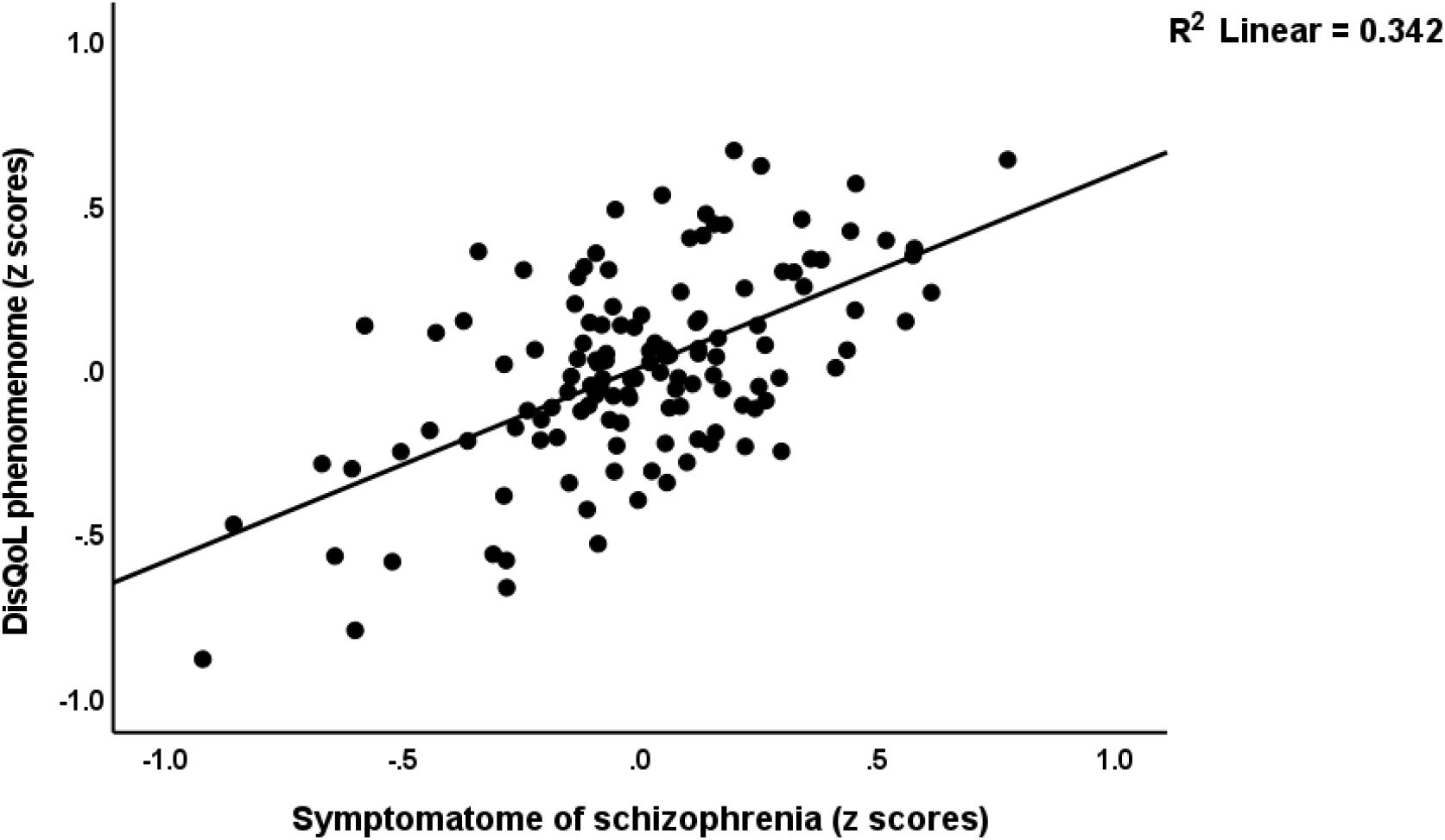
Partial regressions of the DisQol phenomenome, a latent vector extracted from 3 Sheehan disability (Dis) domains and 4 WHO-Quality of Life (QoL) domains on the symptomatome of schizophrenia, a latent vector extracted from all symptom domains, after adjusting for the effects of confounders.

**Figure 3.**
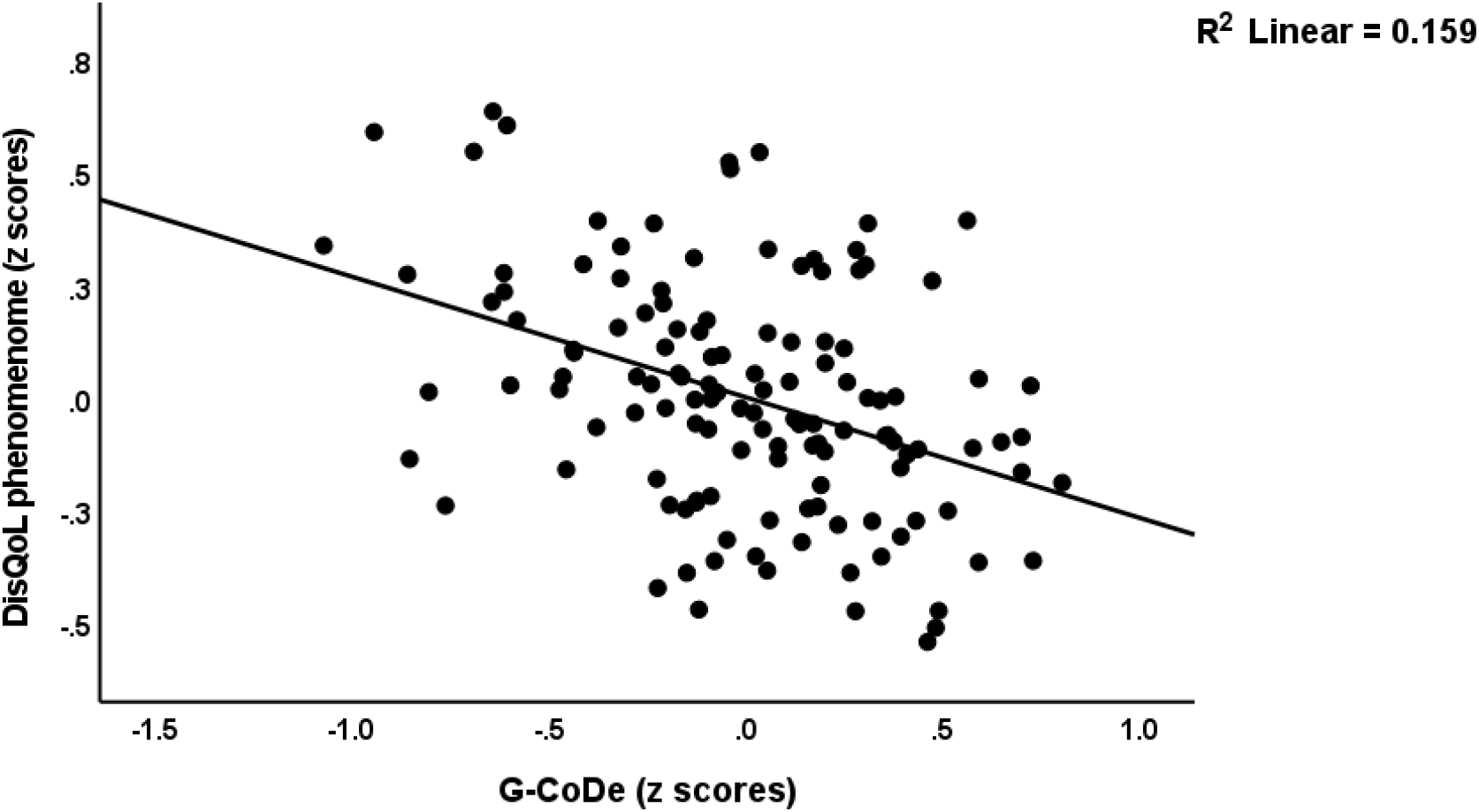
Partial regression of the DisQol phenomenome, a latent vector extracted from 3 Sheehan disability (Dis) domains and 4 WHO-Quality of Life (QoL) domains, on the generalized cognitive decline (G-CoDe) of schizophrenia, a latent vector extracted from cognitive test results, after adjusting for the effects of confounders.

We have also constructed a second PLS model (**Figure 4**) in which we combine all G-CoDe, symptomatome and DisQoL features into one LV and examine the effects of the integrated IL6IL23Th17 index LV on this “worsening LV”. The model quality data and construct reliability validity of the worsening LV were more than adequate with SRMR=0.020, and AVE=0.790; composite reliability, Cronbach alpha > 0.85, all loadings > 0.702, and a construct cross-validated redundancy of 0.492. PLSPredict shows that all Q^2^ predict values were positive indicating that the prediction error was less than the error of the naivest benchmark. Complete compositional invariance was observed using multi-group and predicted-oriented segmentation analysis. Confirmatory Tetrad Analysis showed that the worsening LV was not mis-specified as a reflective model. We found that 61.2% of the variance in the worsening index could be explained by the regression on the IL6IL23Th17 axis LV. **Figure 4** shows the partial regression of the worsening index on the IL6IL23Th17 index in the restricted group of schizophrenia patients (adjusted for age, sex, BMI, education and TUD). Using regression analyses, we did not find any significant effects of the drug state of the patients on the worsening index or the DisQol scores (even without FDR p correction).

**Figure 4.**
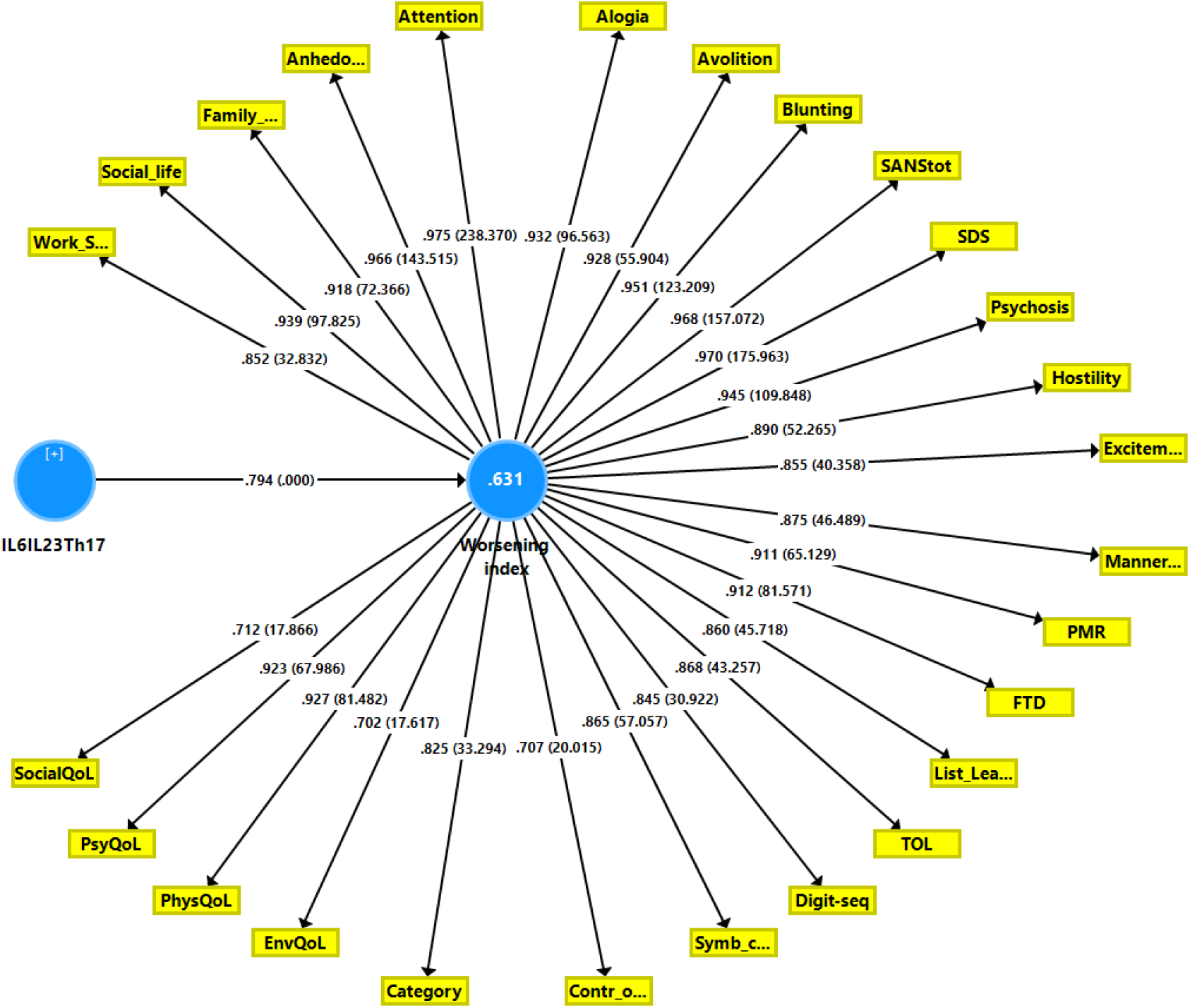
Results of a second Partial Least Squares path analysis. This PLS analysis examines the associations between the IL6IL23Th17 (interleukin, T helper) axis score and the worsening index, a factor extracted from 3 Sheehan disability domains, 4 health-related quality of Life (QoL) data, the symptomatome and the G-CoDe (generalized cognitive decline). See Figure 1 for further explanation. Shown are path coefficients with p value, the factor loadings with p value and explained variance (white figure in the blue circles).

**Figure 5.**
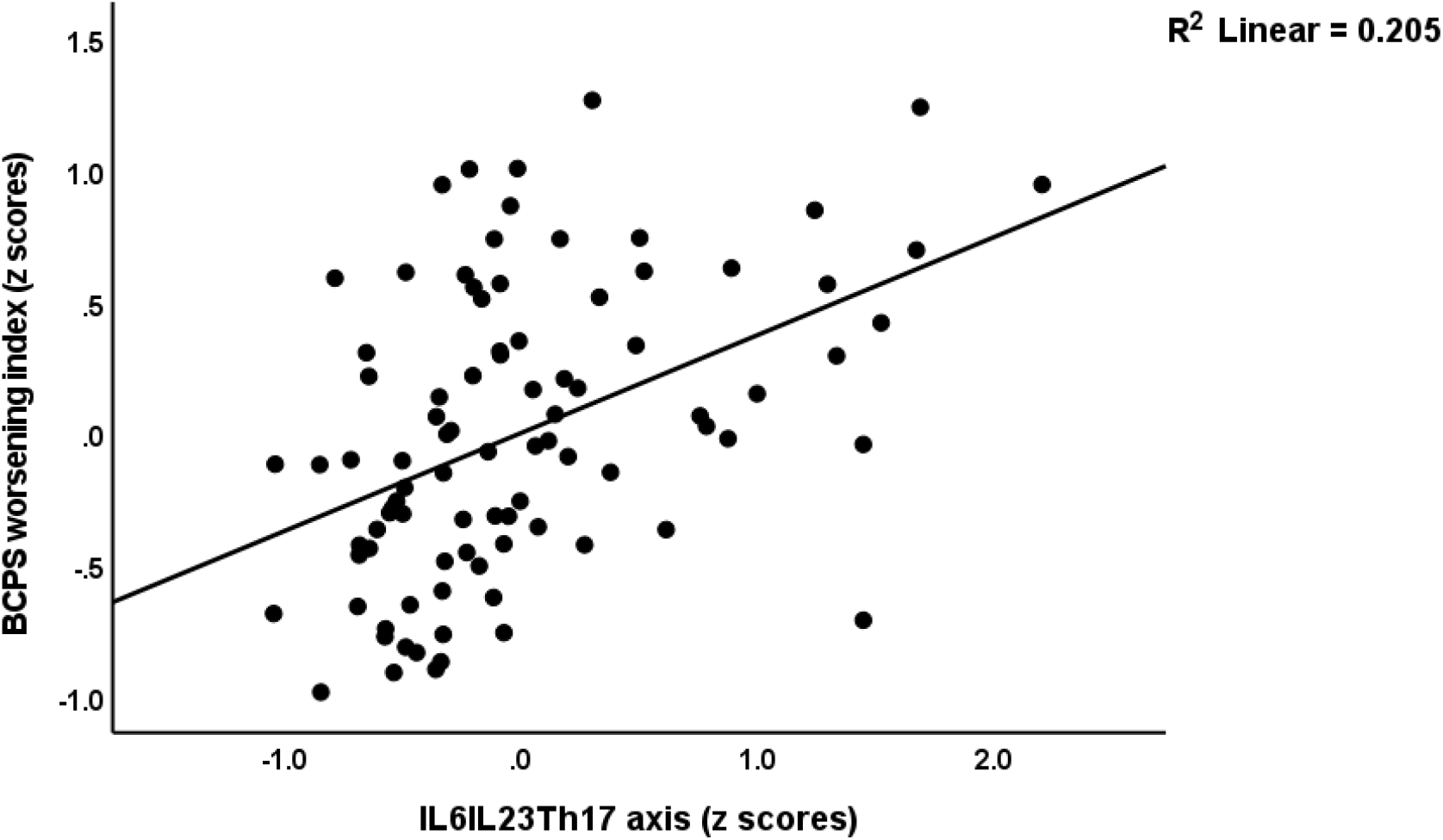
Partial regression of the BCPS (behavior-cognitive-psycho-social) worsening index, a latent vector extracted from 3 Sheehan disability domains and 4 WHO-QoL (Quality of Life) domains, the generalized cognitive decline and the symptomatome on the IL6IL23Th17 (interleukin – T helper) axis severity, computed in the restricted group of schizophrenia patients after adjusting for the effects of confounders.

## Discussion

The first major finding of this study is that lowered HR-QoL and increased disabilities in schizophrenia are indirectly predicted by the IL6IL23Th17 axis and that these effects are mediated by the G-CoDe and symptomatome. These findings extent a previous report that increased levels of IgAs to neurotoxic TRYCATs strongly predict lowered HR-QoL in schizophrenia patients and that these effects are mediated by cognitive deficits and the PHEMN symptoms of schizophrenia (Sirivichayakul et al., 2019; Maes and Kanchanatawan, 2021).

Firstly, the results of the current study corroborate prior research indicating that psychiatric symptoms in general, as well as positive and negative symptoms, may have an effect on HR-QoL in schizophrenia (Galuppi et al., 2010; Sim et al., 2004; Ritsner et al., 2005; Becker et al., 2005; Fitzgerald et al., 2001; Gorna et al., 2014; Savill et al., 2016). As mentioned in the introduction, earlier research often produced inconsistent findings, with some reports indicating that either positive or negative symptoms are the major determinants of lowered HR-QoL (Becker et al., 2005; Fitzgerald et al., 2001; Norman et al., 2000; Savill et al., 2016). Nevertheless, the results of the current study and Maes and Kanchanatawan (2021) show that a general symptomatome factor, rather than positive or negative symptoms, explains lowered HR-QoL and disabilities. Since PHEMN, PMR and FTD symptoms are manifestations of the same underlying core (Almulla et al., 2021; Maes et al., 2019; Maes and Anderson, 2021), it is not adequate to examine whether negative or PHEM symptoms would have a stronger effect. It is also interesting to note that the deficit phenotype is accompanied by lowered HR-QoL and increased disabilities as compared with the non-deficit phenotype (Maes and Kanchanatawan, 2021). In fact, this link may be explained by the fact that the deficit phenotype is associated with a higher severity of the symptomatome and Go-CoDe than nondeficit schizophrenia.

Second, not only the symptomatome but also cognitive deficits such as in episodic and semantic memory, attention, and executive functions have direct effects on the DisQoL phenomenome and additionally indirect effects that are mediated by the symptomatome. These results extend previous reports that cognitive deficits including in verbal memory, attention and speed of information processing, executive dysfunctions and impairments in working memory, verbal ability, verbal list learning, and processing speed, are associated with lowered HR-QoL (Ueoka et al. 2010; Alptekin et al., 2005; Tolman and Kurtz, 2012; Mohamed et al. 2008; Keefe and Harvey, 2012; McGurk and Mueser, 2003; Kanchanatawan et al., 2019). The influence of positive versus negative symptoms versus cognitive impairments on HR-QoL is then explored in some of these papers, as well as whether symptoms contribute more to HR-QoL than cognitive deficits. Nevertheless, all these examinations are not really adequate because one has to consider that part of the effects of cognitive deficits on the phenomenome are mediated by the symptomatome (this study and Maes and Kanchanatawan, 2021). Thus, deficits in learning, executive functions, and working, and semantic and episodic memory might lead to false memory generation and false recall and hence explain in part formal thought disorders, disorganized thought processes and paranoia (Orellana and Slachevsky, 2013; Keefe and Harvey, 2015; Corlett et al., 2007; Maes and Kanchanatawan et al., 2021). Because cognitive impairments which reflect central neurocircuitry dysfunctions often precede the development of acute psychotic episodes, it was argued that cognitive abnormalities contribute to the clinical symptoms of schizophrenia (Harvey et al., 2006; Tamminaga, 2006; Maes and Kanchanatawan, 2021). Verbal memory deficits and attentional impairments, for example, predict psychotic symptoms in ultra-high-risk persons (Brewer et al., 2005; Hawkins et al., 2004). Moreover, in schizophrenia, a single latent trait underpins cognitive deficits, PHEM and negative symptoms indicating that the objective cognitive impairments of the G-CoDe as well as the symptomatome are strongly intertwined indicators reflecting the core of schizophrenia, namely aberrations in “prefronto-striato-thalamic, prefronto-parietal, prefronto-temporal, and dorsolateral prefrontal cortex, as well as hippocampus and amygdala” (Maes and Kanchanatwan et al., 2021; Maes et al., 2019; Orellana et al., 2012; Orellana and Slachevsky, 2013).

Third, the current study’s multistep, multiple mediation PLS model demonstrated that the IL6IL23Th17 axis’ neuroimmunotoxic effects and lowered protection by calcium and magnesium greatly influenced HR-QoL and disabilities, although regression analysis shows that this axis has only minimal impact on the DisQoL phenomenome. Because most researchers do not use multistep mediation models to investigate the impact of biomarkers on the phenomenome, such indirect connections may go undetected, despite their importance. Although a common latent trait underpins IL-6, IL-23, and the Th17-linked cytokines (IL-17, IL-21, IL-22, and TNF-α), we also found that both IL-6 and IL-23 are significantly linked to the Th-17-associated cytokines and, as a result, to the G-CoDe, symptomatome, and, consequently, to HR-QoL and disabilities. As previously discussed, IL-23 and IL-6 are important inducers of proliferation and survival of the pathogenic Th-17 phenotype as well as IL-17 production (Al-Hakeim et al., 2022). Increased IL-6, IL-23, and Th-17-related cytokines have a variety of neuroimmunotoxic effects, including activating autoimmune responses, maintaining persistent peripheral inflammation and neuroinflammation, lowering hippocampal neurogenesis, microglial activation, CNS tissue damage, and induction of the JAK-STAT and MAP-kinase pathways (Nitsch et al., 2019; Tahmasebinia and Pourgholaminejad, 2017; Liu et al., 2014; Langrish et al., 2005; Leavy et al., 2014; Lee et al., 2022; Liang et al., 2010). In schizophrenia, such mechanisms have been linked to abnormalities in neuroplasticity, synapse assembly, synapse structure, axonal branching and axogenesis, excitatory synaptic functioning, and pre- and post-synaptic neural connections (Maes et al., 2021a). Furthermore, the combined negative effects of the IL6IL23Th17 axis and increases in other neuroimmunotoxic consequences such as TRYCATS, oxidative damage, hypernitrosylation, breakdown of the paracellular pathways, and LPS from Gram-negative bacteria may result in enhanced impairments to neuronal functioning in the above-mentioned brain circuits (Maes et al., 2021a; 2021b). Such effects will likely be more prominent when neurotrophic (e.g. brain-derived neurotrophic factor signaling) and antioxidant (e.g. paraoxonase activity) defenses are insufficient (Maes et al., 2021a; 2021b). In this respect, we observed that lower magnesium levels contribute to the G-CoDe, whereas lower calcium levels contribute to the symptomatome. Some authors, although not all, have documented changes in these ions in schizophrenia (Bojarski et al., 2010; Li et al., 2018; Levine et al., 1996; Kirov et al., 1994; Khan et al., 1990; Jamilian, 2011; Kirov and Tsachev, 1990). Reduced calcium and magnesium levels, as well as impaired antioxidant defenses, may be the result of immune-inflammatory processes (Mousa et al., 2021). Magnesium also acts as an antagonist at the glutamatergic NMDA receptor (Sowa-Kucma et al., 2013), while intracellular calcium controls the negative feedback on NMDA channels (Xin et al., 2005), influencing excitotoxicity and neuronal plasticity.

Finally, the current findings were able to confirm prior results that the G-CoDe, symptomatome, and phenomenome of schizophrenia are all underpinned by a latent vector, namely the BCPS worsening index (Maes et al., 2021b). These components, by inference, are indications of a similar core, namely BCPS worsening, which is the result of neuroimmunotoxic effects of activated peripheral immune-inflammatory pathways on neural processes in particular brain circuits. While the pathogenic IL6IL23Th17 axis was shown to be a major predictor of this BCPS worsening index in the current investigation, a prior publication found that this index was linked to various neuroimmunotoxic pathways, including LPS, nitro-oxidative pathways, and other neurotoxic cytokines (Maes et al., 2021b).

## Conclusions

A significant portion of the DisQoL phenome is predicted by the G-CoDe and the symptomatome, while the IL6IL23Th117 axis has no significant effect beyond and above these clinical features. Nevertheless, mediation analysis shows that the IL6IL23Th17 axis index has highly significant indirect effects on the DisQoL phenome that are mediated by the G-CoDe and the symptomatome. Moreover, the IL6IL23Th17 axis accounted for 63.1 percent of the variation in the BCPS worsening index, namely a latent variable derived from G-CoDe, symptomatome, HR-QoL, and disability data. The neuroimmunotoxi effects of the IL6IL23Th17 axis not only impact G-CoDe and the symptomatome, but also the different domains of the DisQoL phenomenome.

## Data Availability

The dataset generated during and/or analyzed during the current study will be available from the corresponding author (MM) upon reasonable request and once the dataset has been fully exploited by the authors.

## Acknowledgment

We acknowledge the staff of Al-Hakeem General Hospital, Psychiatric Unit in Najaf Governorate, for their help in collecting samples. We also acknowledge the work of the high-skilled staff of Asia Clinical *L*aboratory in Najaf city for their help with the ELISA assays.

## Author’s contributions

All the contributing authors have participated in preparation of the manuscript.

*Compliance with ethical standards*

## Research involving human participants

Approval for the study was obtained from the Institutional Review Board of the University of the University of Kufa, Iraq (82/2020)

## Informed consent

All participants and first-degree relatives of participants with schizophrenia gave written informed consent before participation in our study.

Data Access Statement.

